# “Super-Papers” Make More Impact: A Bibliometric and Altmetric Databases Analysis of MASLD Publications

**DOI:** 10.1101/2025.07.23.25332071

**Authors:** Tal Kaminski-Rosenberg, Jeffrey V Lazarus, Shira Zelber-Sagi

**Affiliations:** School of Public Health, Faculty of Social Welfare and Health Sciences, University of Haifa, Haifa, Israel; Medical Library, Rambam Health Care Campus and Technion Faculty of Medicine, Haifa, Israel; City University of New York Graduate School of Public Health and Health Policy (CUNY SPH), New York, NY, USA; Barcelona Institute for Global Health (ISGlobal), Barcelona, Spain; Facultat de Medicina i Ciències de la Salut, Universitat de Barcelona (UB), Barcelona, Spain

**Keywords:** fatty liver, healthy lifestyle, policy citations, research impact, steatohepatitis

## Abstract

**Aims:** Traditional evaluation of research impact was based on citation analyses, but the alternative metrics (altmetrics) measure broader societal impact. We aimed to explore the association between academic and societal impact of papers across two Metabolic dysfunction-associated steatotic liver disease (MASLD) fields; lifestyle and burden/policy, and to identify the factors predicting the citation of a paper within policy documents.

**Methods:** We conducted a database analytic study of the 100 top-cited and 100 top-Altmetric Attention Score (AAS) papers dealing with MASLD in relation to lifestyle or to burden/policy fields. The papers were retrieved from the Web of Science platform and Altmetric Explorer database. Policy citation data were complemented from the Overton database. Papers within both 100 top-AAS and 100 top-cited papers lists in each field were defined as “super-papers”.

**Results:** A moderate correlation was found between citations and AAS in both lifestyle and burden/policy fields (r=0.399, P<0.001 and r=0.455, P<0.001, respectively). A five-unit increase in the journal’s impact factor augmented the probability of a lifestyle paper being a “super-paper” by 10% (95% confidence interval 1.02-1.19). Open access publishing increased the likelihood of a burden/policy paper being a “super-paper” by 3.33-fold (1.26-8.85, P=0.015). “Super-papers” were significantly more likely to be cited within policy documents compared to being either top-cited or top-AAS papers by 2.60 (1.06-6.37 P=0.036) and 4.53-fold (1.68-12.19 P=0.003) respectively in the burden/policy field.

**Conclusion:** Papers in the field of MASLD that are better disseminated across both academic and societal platforms are associated with higher policy impact.

## Introduction

Measuring the impact of research is of great interest to all key stakeholders, including researchers, institutions, funders and the public. Since the 1960s, the metrics used to do this have been based on citation analyses, which only show the academic impact of research papers and overlook their societal influence(1, 2). The advancement of digital technologies allows for the measurement of the impact of research papers at a broader scale via alternative metrics, or altmetrics; these enable capturing a paper’s influence in the “real world” and in real-time, via many online sources(3).

Metabolic dysfunction-associated steatotic liver disease (MASLD), formerly non-alcoholic fatty liver disease (NAFLD)(4), is the most common liver disease globally, with an estimated overall prevalence among adults of 32%(5). MASLD is closely related to obesity and type 2 diabetes, with estimates of the prevalence of MASLD among people living with these conditions being as high as double its global prevalence(6).

For the past decades, the first-line treatment of MASLD has been a healthy diet and lifestyle (7, 8). Only as of 2024, the first drug to treat the more advanced form of MASLD, received approval by the United States Food and Drug Administration, for use alongside diet and exercise(9).

Despite the high prevalence, the academic and societal impact of MASLD publications has not been examined extensively. Previous studies on citations and altmetrics show such data to be field-specific, preventing the extrapolation of findings from one field to another(10). This study thus aimed to: (1) explore the academic and societal dissemination and impact of papers across two MASLD fields of study (1a) lifestyle for the prevention and treatment of MASLD (named lifestyle field) and (1b) the human and economic burden, awareness and policies for MASLD (named burden/policy field). (2) determine the relationship between the academic and societal impact within each field and (3) identify the factors predicting the citation of a paper within policy documents.

## Methods

We conducted a descriptive and analytical database study of bibliometric and altmetric data of papers in the lifestyle and burden/policy fields in MASLD. A sample size of 100 papers for each field and metric (total n=400) was determined based on standard statistical power calculations and the acceptable sample size found in similar studies on medical research(11, 12).

### Citations and Journal Impact Factor data collection

The Web of Science (WoS) platform (Clarivate, https://www.webofscience.com) was used to search for the highest cited papers in both fields and extract the citations, number of authors and affiliations and first author’s regional data of all analyzed papers. WoS is considered the gold standard for citation analysis(13) and its coverage of medical literature is high(14), enhancing the accuracy and reliability of the data obtained.

The Journal Citation Report (JCR) database (Clarivate, https://jcr.clarivate.com) was the source for the Journal Impact Factor (JIF) and its category and quartile data. JCR is the official source for this data.

### Altmetric data collection

The Altmetric Explorer (AE) database (https://www.altmetric.com/explorer) was used to search for papers with the highest Altmetric Attention Scores (AAS) in both fields and to extract Altmetric data of all papers in the study. AAS is an automatically calculated weighted count of all of the attention a research output has received online, in sources tracked by the Altmetric company (15). The data for top-AAS papers were downloaded from the AE database, along with search results. Altmetric data for top-cited papers were searched for within the AE database via their digital object identifier (DOI) number. Altmetric’s tools are accepted and used in more than 50% of studies within the field of altmetrics(16).

### Policy citations data collection

Policy document citation data were extracted from the Overton database (https://app.overton.io) via the paper’s DOI, in addition to the Altmetric policy citation data. Duplicate findings were documented and removed. Founded in 2019, the Overton database has a high coverage of organizations, countries, languages and policy document types (including governmental and intergovernmental guidance, think-tank research, working papers, reports and clinical guidelines). Medicine is among the best-covered disciplines within the database(17).

### Search and screening process

To identify the papers for analysis, comprehensive search strategies were developed for the WoS platform and AE database. The full search strategies can be found in Supplemental online materials 1-4. Before screening, search results were sorted by “citations highest first” (in WoS) or “AAS highest first” (in AE). The screening process was done by KRT according to the inclusion and exclusion criteria. To be included, papers had to: (1) be about humans and published since 1 January 2014; (2) be research articles, reviews, systematic reviews and meta-analyses, editorials, clinical practice guidelines (CPGs, which include recommendations related to diet or physical activity) or comments; and (3) have as their main subject the association of MASLD with diet and physical activity or the human and economic burden of, policies for and awareness around MASLD. Publications were excluded if they were: (1) letters to the editor/correspondence, case reports, preprints, conference abstracts and dissertations; (2) on animal and basic research; (3) on herbal and dietary supplements, sleep, smoking and alcohol consumption; and (4) in journals not indexed in the 2022 edition of the JCR core collection (and thus had no impact factor). Uncertainties during the screening process were resolved in consultation with SZS and JVL.

### Variables extracted for paper characterization

The variables collected were categorized as paper’s level variables, author’s level variables and journal’s level variables. A full description of the variables can be found in **Appendix I**. Paper’s level variables included its status (i.e. top-cited, top-AAS, or “super-paper” - papers within both 100 top-AAS and 100 top-cited papers lists in each field), AAS, publication type (i.e. research article, review, systematic review and meta-analysis or other (grouping editorials, CPGs and comments which were few), open access (OA) status and the number of citations, years since publication, citations within policy documents, patents and Twitter (X), Mendeley reader, news, blogs, Facebook, Wikipedia and YouTube mentions.

Author level variables included the number of authors and affiliations and first author region (i.e., North and South America (including Canada), Australia and New Zealand, Europe, Asia and the Middle East, there were no papers from Africa). Journal level variables included the JIF category as per the JCR database (i.e., gastroenterology and hepatology, nutrition and dietetics, endocrinology and metabolism or other) and JIF quartile (Q) within the JCR category (i.e., 1, 2 or 3/4).

### Data analysis

All analyses were undertaken via the Statistical Package for Social Sciences software, version 27.0 (IBM). All continuous variables were not normally distributed according to the Kolmogorov-Smirnov test. For descriptive statistics, categorical variables are presented as n (%) and continuous variables as a median and interquartile range (IQR). Categorical variables were compared between groups using the chi-square or Fisher test, as appropriate. Continuous variables were compared using the Mann-Whitney test between two groups or the Kruskal-Wallis test among three groups. Spearman’s correlation coefficient (r_s_) was calculated to assess the correlation between the number of citations and AAS in both fields. Multivariable logistic regression models were used to test the probability of a paper being a “super-paper” and the likelihood of a paper being cited within a policy document. Results are presented as odds Ratio (OR) and 95% confidence interval (CI). Factors and covariates within the models were those that are independent of the “super-paper” status (excluding the number of citations, AAS and its components: mentions in X (Twitter), Facebook, Wikipedia, Blogs, YouTube, news, patents) and significant, or borderline-significant, according to the univariable analysis. Policy citations were used in the models since they were mostly (80%) derived from the Overton database and were not part of the AAS calculation made by Altmetric (**Appendix II**).A P-value of <0.05 was considered statistically significant for all analyses.

Artificial intelligence (AI) tools were used in this work for the de-duplication of affiliations and papers, and for help in paraphrasing the authorś own ideas. All information regarding said use can be found in **Appendix III**.

## Results

### Lifestyle papers search results and screening process

The WoS platform was searched on 15 January 2024 to identify the top-cited lifestyle papers. The search results and screening process are shown in **Figure 1**. The full list of the 100 top-cited lifestyle papers is available in **Supplemental online material 5**. The Altmetric Explorer (AE) database was searched on 15 January 2024 to identify the top-AAS lifestyle papers. The search results and screening process are depicted in **Figure 1**. The full list of the 100 top-AAS lifestyle papers is available in **Supplemental online material 6**.

**Figure 1.**
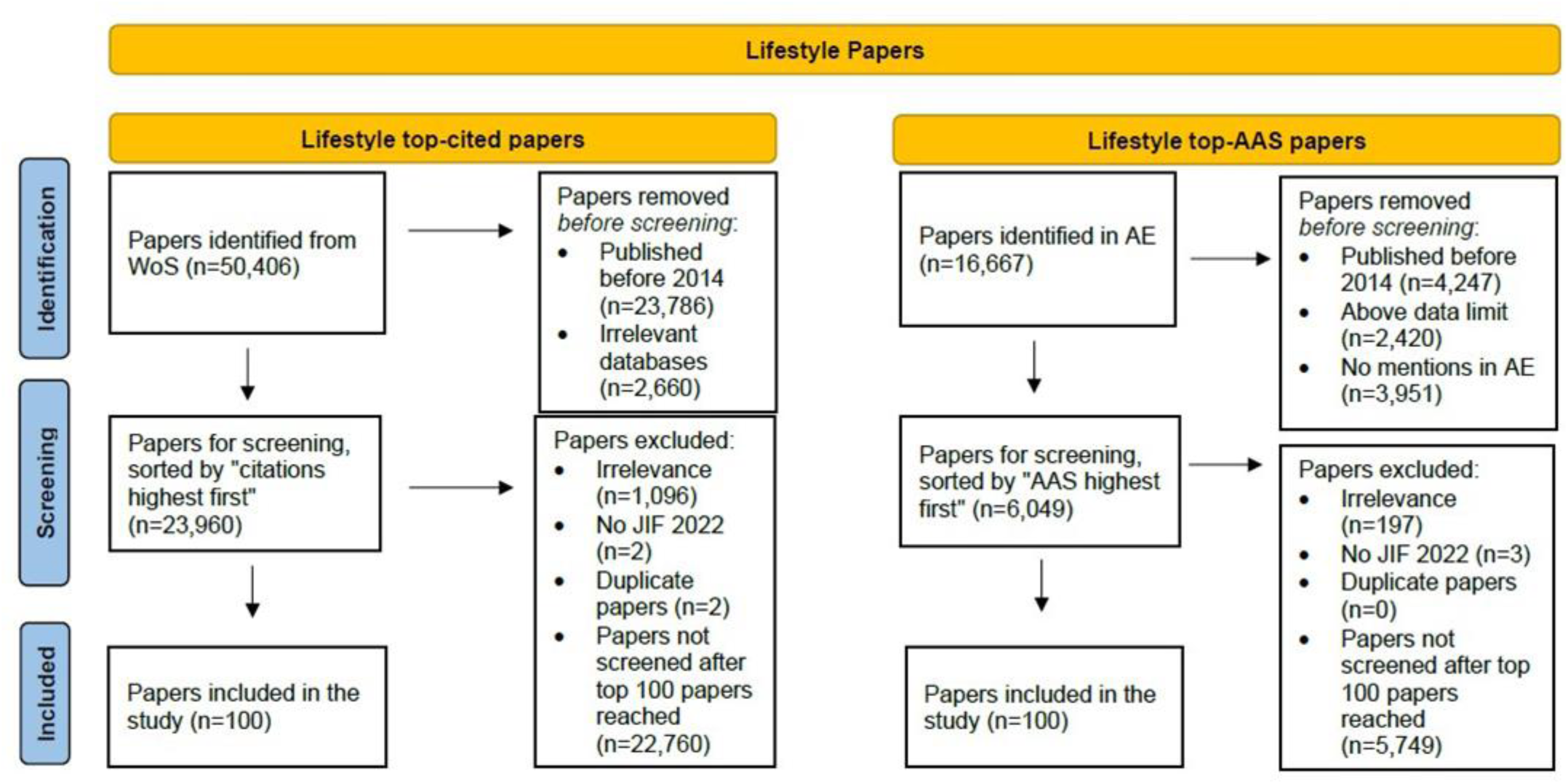
Search results and screening process of lifestyle papers. Adapted from the Preferred Reporting Items for Systematic reviews and Meta-Analyses flow diagram found at https://www.prisma-statement.org/prisma-2020-flow-diagram Abbreviations: AAS, Altmetric Attention Score; AE, Altmetric Explorer; JIF, Journal Impact Factor; WoS, Web of Science.

### Burden/policy papers search results and screening process

The WoS platform was searched on 7 February 2024 to identify the top-cited burden/policy papers. The search results and screening process are shown in **Figure 2**. The full list of the 100 top-cited burden/policy papers is available in **Supplemental online material 7**. The AE database was searched on 7 February 2024 to identify the top-AAS burden/policy papers. The search results and screening process are depicted in **Figure 2**. The full list of the 100 top-AAS burden/policy papers is available in **Supplemental online material 8**.

**Figure 2.**
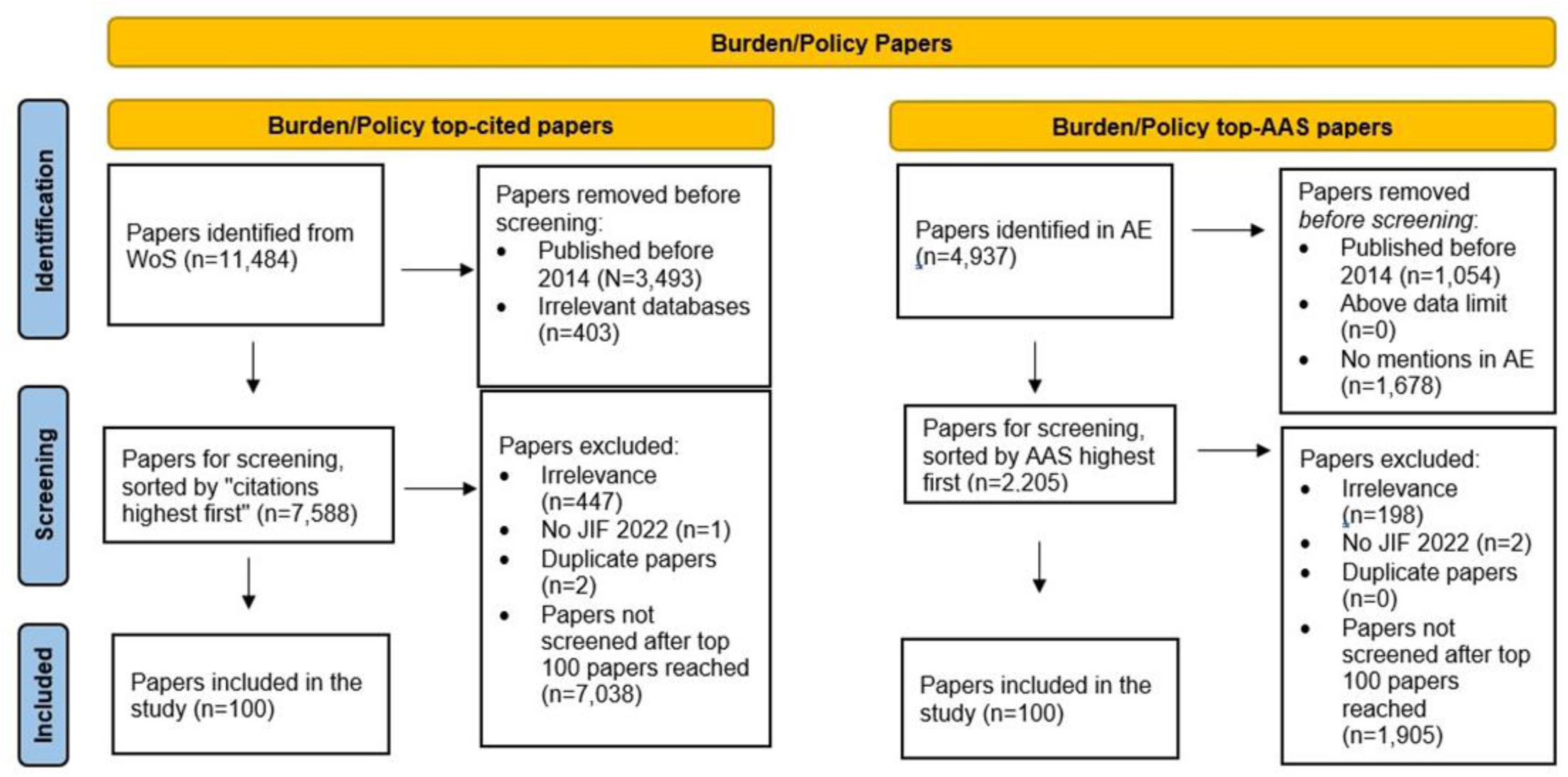
Search results and screening process of burden/policy papers. Adapted from the Preferred Reporting Items for Systematic reviews and Meta-Analyses flow diagram found at https://www.prisma-statement.org/prisma-2020-flow-diagram Abbreviations: AAS, Altmetric Attention Score; AE, Altmetric Explorer; JIF, Journal Impact Factor; WoS, Web of Science.

### Correlation between the number of citations and Altmetric Attention Score

The r_s_ between the number of citations and AAS for the top-cited lifestyle papers was 0.399 (P<o.oo1), indicating a moderate correlation. The r_s_ between the number of citations and AAS for the top-cited burden/policy papers was 0.455 (P<o.oo1), indicating a moderate correlation.

### “Super-paper” analysis

Overlapping publications between the lists of 100 top-cited papers and 100 top-AAS papers were detected and defined as “super-papers”. There were 26 duplicate papers in the lifestyle field and 41 in the burden/policy field.

#### Lifestyle “super-papers”

After de-duplication, the total number of lifestyle papers was 174: 74 top-cited papers (n=74), 74 top-AAS papers (n=74), and 26 “super-papers” (**Table 1**). The median number of years since publication was 3 (IQR 3), 8 (IQR 2) and 6 (IQR 2) (P<0.001) for the top-AAS, top-cited and “super-papers”, respectively. At 25.1 (IQR 18.4), the JIF median of the “super-papers” was significantly higher than that of the top-cited and top-AAS papers (P<0.001), whose median did not differ, at 5.9 (IQR 4.75) and 6 (IQR 7.9), respectively. The number of mentions in Wikipedia, Facebook and YouTube of the “super-papers” was significantly higher than said counts for top-AAS and top-cited papers (these variables were not included in the multivariable analysis, since they are part of the AAS score).

**Table 1.** Comparison of lifestyle top-cited, top Altmetric Attention Score and “super-papers” characteristics.

| Variable | Lifestyle top-cited papers (n=74) | Lifestyle top-AAS papers (n=74) | Lifestyle "Super-papers" (n=26) | P Value |
| --- | --- | --- | --- | --- |
| **Paper and Journal characteristics** |  |  |  |  |
| OA Status<sup>1</sup> | 51(68.9) | 60(81.1) | 22 (84.6) | 0.149 |
| Publication Type* |  |  |  | 0.092 |
| Research Article | 32(43.2) | 47(63.5) | 15 (57.7) |  |
| Review | 29(39.2) | 13(17.6) | 8 (30.8) |  |
| SR/MA | 8(10.8) | 9(12.2) | 1 (3.8) |  |
| Other | 5(6.8) | 5(6.8) | 2(7.7) |  |
| JIF Category in JCR* |  |  |  | 0.400 |
| Gastroenterology and Hepatology | 38(51.4) | 29(39.2) | 15 (57.7) |  |
| Nutrition and Dietetics | 17(23) | 23(31.1) | 4 (15.4) |  |
| Endocrinology and Metabolism | 7(9.5) | 10(13.5) | 1 (3.8) |  |
| Other | 12(16.2) | 12(16.2) | 6 (23.1) |  |
| Q of JIF in JCR* |  |  |  | 0.068 |
| 1 | 48(64.9) | 52(70.3) | 24 (92.3) |  |
| 2 | 19(25.7) | 18(24.3) | 1 (3.8) |  |
| 3/4 | 7(9.5) | 4(5.4) | 1 (3.8) |  |
| First author region* |  |  |  | 0.242 |
| America, Canada, Australia, NZ | 25(33.8) | 29(39.2) | 8(30.8) |  |
| Europe | 26(35.1) | 33(44.6) | 13 (50) |  |
| Asia, Middle East | 23(31.1) | 12(16.2) | 5(19.2) |  |
| Years since publication | 8(2)<sup>a</sup> | 3(3)<sup>b</sup> | 6(2)<sup>c</sup> | <0.001 |
| JIF (2022)** | 5.9(4.75)<sup>a</sup> | 6(7.9)<sup>a</sup> | 25.1(18.4)<sup>b</sup> | <0.001 |
| No. of authors** | 6(7)<sup>a</sup> | 9(9)<sup>b</sup> | 8.5(9)<sup>a,b</sup> | 0.017 |
| No. of affiliations** | 3(2) | 3(3) | 4 (5) | 0.099 |
| **Components of the impact measures** |  |  |  |  |
| No. of citations** | 112 (58)<sup>a</sup> | 19(38)<sup>b</sup> | 127.5 (84)<sup>a</sup> | <0.001 |
| No. of Mendeley readers** | 212(105)<sup>a</sup> | 53.5(76)<sup>b</sup> | 237(183)<sup>a</sup> | <0.001 |
| AAS** | 25(41)<sup>a</sup> | 114.5(175)<sup>b</sup> | 246.5(452)<sup>b</sup> | <0.001 |
| No. of Twitter(X) mentions** | 10(43)<sup>a</sup> | 72(146)<sup>b</sup> | 171.5 (427)<sup>b</sup> | <0.001 |
| No. of news mentions** | 1(4)<sup>a</sup> | 9(19)<sup>b</sup> | 9(37)<sup>b</sup> | <0.001 |
| No. of policy mentions** | 0(1)<sup>a</sup> | 0(0)<sup>b</sup> | 0(1)<sup>a</sup> | <0.001 |
| No. of blog mentions** | 0(1)<sup>a</sup> | 0(1)<sup>a,b</sup> | 1(2)<sup>b</sup> | 0.016 |
| No. of patent mentions** | 0(0)<sup>a</sup> | 0(1)<sup>b</sup> | 0(0)<sup>a</sup> | 0.022*** |
| No. of Facebook mentions** | 0(2)<sup>a</sup> | 1(2)<sup>a</sup> | 5(10)<sup>b</sup> | <0.001 |
| No. of Wikipedia mentions** | 0(0)<sup>a</sup> | 0(0)<sup>a</sup> | 0(1)<sup>b</sup> | <0.001 |
| No. of YouTube mentions** | 0(1)<sup>a</sup> | 0(1)<sup>a</sup> | 2(5)<sup>b</sup> | 0.002 |

\*n(%); \*\* Median (IQR); \*\*\* Not Significant after Bonferroni adjustment for multiple tests; a,b,c: Shows the statistically significant differences between the 3 groups by Kruskal-Wallis test. Abbreviations: AAS: Altmetric Attention Score; JCR: Journal Citation Report (Clarivate); JIF: Journal Impact Factor; No.: Number; OA: Open Access; Q: Quartile; SR/MA: Systematic Review/Meta-Analysis. Definitions: "Super-papers"-duplicate articles appearing in both 100 top-cited papers and 100 top-AAS papers.

In a multivariable analysis including the number of years since publication and JIF (**Table 2**) a significant association was identified between the JIF and “super-paper” status (OR=1.02; 95% CI 1.01-1.04, P=0.019, per one unit increase). A five-unit increase in JIF augmented the probability of a paper being a “super-paper” by 1.1 (95% CI 1.02-1.19). The number of years since publication were not significantly related to the likelihood of a paper being classified as a “super-paper”.

**Table 2.** Multivariable analysis of the association between years since publication and Journal Impact Factor on “super-paper” status.

| Variables | "Super-papers" status |  |
| --- | --- | --- |
|  | OR (95% CI) | P value |
| Years since publication | 1.07(0.91-1.25) | 0.396 |
| **JIF (2022)** | **1.02(1.01-1.04)** | **0.019** |

Abbreviations: JIF: Journal Impact Factor. "Super-papers" definition: duplicate articles appearing in both 100 top-cited papers and 100 top-AAS papers.

#### Burden/policy “super-papers”

Excluding duplicates, the total number of burden/policy papers was 159: top-cited papers (n=59), top-AAS papers (n=59), and “super-papers” (n=41) (**Table 3**) OA status differed significantly across the three paper categories (P=0.005), with top-cited papers had the lowest percentage of OA papers (52.5%), while the “super-papers” had the highest (82.9%). The median years since publication for the top-AAS papers was 3 years (IQR 4), significantly lower than the other two groups which had about two-fold greater median (P<0.001).

**Table 3.** Comparison of burden/policy top-cited, top Altmetric Attention Score and “super-papers” characteristics.

| Variable | Burden/policy top-cited papers n=59 | Burden/policy top-AAS papers n=59 | Burden/policy "Super-papers" n=41 | P-value |
| --- | --- | --- | --- | --- |
| **Paper and Journal characteristics** |  |  |  |  |
| OA Status* | 31(52.5) | 41(69.5) | 34(82.9) | 0.005 |
| Publication Type* |  |  |  | 0.215 |
| Research Article | 28(47.5) | 34(57.6) | 23(56.1) |  |
| Review | 13(22) | 10(16.9) | 9(22) |  |
| SR/MA | 17(28.8) | 10(16.9) | 5(12.2) |  |
| Other | 1(1.7) | 5(8.5) | 4(9.8) |  |
| JIF Category in JCR* |  |  |  | 0.610 |
| Gastroenterology and Hepatology | 45(76.3) | 45(76.3) | 35 (85.4) |  |
| Nutrition and Dietetics | 0 | 1(1.7) | 0 |  |
| Endocrinology and Metabolism | 6(10.2) | 3(5.1) | 1(2.4) |  |
| Other | 8(13.6) | 10(16.9) | 5(12.2) |  |
| Q of IF in JCR* |  |  |  | 0.021 |
| 1 | 36(61) | 48(81.4) | 35(85.4) |  |
| 2 | 15(25.4) | 5(8.5) | 5(12) |  |
| 3/4 | 8(13.6) | 6(10.2) | 1(2.4) |  |
| First author region* |  |  |  | 0.028 |
| America, Canada, Australia, NZ | 25(42.4) | 31(52.5) | 26(63.4) |  |
| Europe | 14(23.7) | 13(22) | 12(29.3) |  |
| Asia, Middle East | 20(33.9) | 15(25.4) | 3(7.3) |  |
| Years since publication** | 6(5)<sup>a</sup> | 3(4)<sup>b</sup> | 5(4)<sup>a</sup> | <0.001 |
| JIF (2022)** | 6.2(8.5)<sup>a</sup> | 13.8(19)<sup>b</sup> | 14(18.5)<sup>b</sup> | <0.001 |
| No. of authors** | 7(6) | 9(7) | 7 (8) | 0.298 |
| No. of affiliations** | 3(3) | 4(6) | 3 (5) | 0.056 |
| **Components of the impact measures** |  |  |  |  |
| No. of citations** | 91(111)<sup>a</sup> | 38(54)<sup>b</sup> | 164 (405)<sup>c</sup> | <0.001 |
| No. of Mendeley readers** | 115(124)<sup>a</sup> | 49(97)<sup>b</sup> | 174 (429)<sup>c</sup> | <0.001 |
| AAS** | 8(14)<sup>a</sup> | 38(47)<sup>b</sup> | 57(131)<sup>b</sup> | <0.001 |
| No. of Twitter(X) mentions** | 2(12)<sup>a</sup> | 40(77)<sup>b</sup> | 38(63)<sup>b</sup> | <0.001 |
| No. of news mentions** | 0(2)<sup>a</sup> | 2(4)<sup>b</sup> | 5(14)<sup>b</sup> | <0.001 |
| No. of policy mentions** | 0(1)<sup>a</sup> | 0(0)<sup>a</sup> | 1(2)<sup>b</sup> | <0.001 |
| No. of blog mentions** | 0(0)<sup>a</sup> | 0(0)<sup>a,b</sup> | 0(1)<sup>b</sup> | 0.013 |
| No. of patent mentions** | 0(0)<sup>a</sup> | 0(0)<sup>a</sup> | 0(1)<sup>b</sup> | <0.001 |
| No. of Facebook mentions** | 0(1)<sup>a</sup> | 0(1)<sup>a,b</sup> | 1 (2)<sup>b</sup> | <0.001 |
| No. of Wikipedia mentions** | 0(0)<sup>a</sup> | 0(0)<sup>a</sup> | 0(0)<sup>b</sup> | 0.001 |
| No. of YouTube mentions** | 0(0)<sup>a</sup> | 0(0)<sup>a</sup> | 0(0)<sup>b</sup> | 0.004 |

\*n(%); \*\* Median (IQR); a,b,c: Shows the specific statistically significant differences between the 3 groups by Kruskal-Wallis test. Abbreviations: AAS: Altmetric Attention Score; JCR: Journal Citation Report (Clarivate); JIF: Journal Impact Factor; No.: Number; OA: Open Access; Q: Quartile; SR/MA: Systematic Review/Meta-Analysis. Definitions: "Super-papers"-duplicate articles appearing in both 100 top-cited papers and 100 top-AAS papers.

The JIF was lowest for the top-cited papers when compared to the other two papers categories (P<0.001) but was not different between the top-AAS papers and the “super-papers”. The number of Wikipedia, patent, policy document and YouTube mentions was significantly higher for the “super-papers”, when compared to the number of mentions of the top-AAS and top-cited papers (these variables were not included in the multivariable analysis since they are part of the AAS score).

In a multivariable analysis (**Table 4**) including the number of years since publication and affiliations, JIF, first author region and Q of JIF in the JCR database, being OA increased the likelihood of a paper being classified as a “super-paper” (OR=3.33; 95% CI 1.26-8.85, P=0.015). All other variables included in the model were non-significant.

**Table 4.** Multivariable analysis of the association between burden/policy papers characteristics and “super-paper” status.

| Variables | "Super-paper" status |  |
| --- | --- | --- |
|  | OR (95% CI) | P value |
| Years since publication | 1.17(0.99--1.37) | 0.053 |
| JIF(2022) | 1.02(0.99-1.05) | 0.131 |
| Number of affiliations | 1.01(0.95-1.07) | 0.779 |
| **OA status** | **3.33(1.26-8.85)** | **0.015** |
| First author region |  | 0.132 |
| Asia, Middle East | Ref(1) |  |
| America, Canada, Australia, NZ | 3.79(1.01-14.26) | 0.049 |
| Europe | 2.65(0.62-11.39) | 0.191 |
| Q of JIF in JCR |  |  |
| 3/4 | Ref(1) | 0.317 |
| 1 | 4.84(0.55-42.84) | 0.156 |
| 2 | 3.02(0.28-30.67) | 0.350 |

Abbreviations: JCR: Journal Citation Report (Clarivate); JIF: Journal Impact Factor; OA: Open Access; Q: Quartile. Definitions: "Super-papers"-duplicate articles appearing in both 100 top-cited papers and 100 top-AAS papers.

### Policy document citation analysis

#### Lifestyle papers

A total of 58 lifestyle papers (33.3%) were cited at least once within a policy document. A comparison of the characteristics between the papers that were cited within policy documents versus those that were not depicted in **Table 5**. Papers with policy citations had significantly higher years since publication, authors and the number of affiliations, JIF, and a larger proportion of “super-papers”. The number of patents, Wikipedia, Facebook and YouTube mentions in the papers with policy citations was significantly higher than the number of mentions in the papers without policy citations.

**Table 5.** The characteristics of lifestyle top Altmetric Attention Score or top-cited papers with and without policy citations.

| Variables | Papers without policy citations<br>n= 116 | Papers with policy citations<br>n=58 | P Value |
| --- | --- | --- | --- |
| **Paper's and Journal's characteristics** |  |  |  |
| OA Status* | 89(76.7) | 44(75.9) | 1 |
| Publication Type* |  |  | 0.065 |
| Research Article | 60(51.7) | 34(58.6) |  |
| Review | 40(34.5) | 10(17.2) |  |
| SR/MA | 10(8.6) | 8(13.8) |  |
| Other | 6(5.2) | 6(10.3) |  |
| JIF Category in JCR* |  |  | 0.449 |
| Gastroenterology and Hepatology | 50(43.1) | 32(55.2) |  |
| Nutrition and Dietetics | 30(25.9) | 14(24.1) |  |
| Endocrinology and Metabolism | 14(12.1) | 4(6.9) |  |
| Other | 22(19) | 8(13.8) |  |
| Q of JIF in JCR* |  |  | 0.349 |
| 1 | 79(68.1) | 45(77.6) |  |
| 2 | 29(25) | 9(15.5) |  |
| 3/4 | 8(6.9) | 4(6.9) |  |
| First author region* |  |  | 0.915 |
| America, Canada, Australia, NZ | 40(34.5) | 22(37.9) |  |
| Europe | 49(42.2) | 23(39.7) |  |
| Asia, Middle East | 27(23.3) | 13(22.4) |  |
| Years since publication** | 4(5) | 7(3)<sup>a</sup> | <0.001 |
| JIF (2022)** | 5.95(5.5) | 7.1(19.9) | 0.050 |
| No. of authors** | 7(7) | 9(9) | 0.026 |
| No. of affiliations** | 3(2) | 4(4) | 0.015 |
| Paper's status* |  |  | 0.001 |
| Top-cited papers | 41(35.3) | 33(56.9) |  |
| Top-AAS papers | 61(52.6) | 13(22.4) |  |
| "Super-papers" | 14(12.1) | 12(20.7) |  |
| **Components of the impact measures** |  |  |  |
| No. of citations** | 73(89) | 121.5(101) | <0.001 |
| No. of Mendeley readers** | 110.5(176) | 220(101) | <0.001 |
| AAS** | 75.5(151) | 85(284) | 0.702 |

| Variables | Papers without<br>policy citations<br>n= 116 | Papers with<br>policy citations<br>n=58 | P<br>Value |
| --- | --- | --- | --- |
| No. of Twitter(X)<br>mentions** | 50(155) | 50(117) | 0.488 |
| No. of news mentions** | 4(13) | 4(22) | 0.501 |
| No. of blog mentions** | 0(1) | 0(1) | 0.112 |
| No. of Patent<br>mentions** | 0(0) | 0(0) | 0.001 |
| No. of Facebook<br>mentions** | 1(2) | 2(7) | 0.003 |
| No. of Wikipedia<br>mentions** | 0(0) | 0(0) | 0.003 |
| No. of YouTube<br>mentions** | 0(1) | 1(3) | 0.012 |

\*n(%); \*\* Median (IQR); Abbreviations: AAS: Altmetric Attention Score; JCR: Journal Citation Report (Clarivate); JIF: Journal Impact Factor; No.: Number; OA: Open Access; Q: Quartile; SR/MA: Systematic Review/Meta-Analysis.

In multivariable analysis (**Table 6**) including all variables that were significantly or borderline-significantly different between papers with and without policy citation on the univariable analysis (**Table 5**), the significant predictors for policy citation were: years since publication (OR=1.53; 95% CI 1.22-1.91, P<0.001, per one-year increase) and affiliations (OR=1.14; 95% CI 1.01-1.28, P=0.029, per an increase in one affiliation) and being a research article, as opposed to review (OR=2.87; 95% CI 1.12-7.4, P=0.029). Being a “super-paper” was not a statistically significant predictor for being cited in policy documents.

**Table 6.** Multivariable analysis of the association between lifestyle paper’s characteristics and having policy citations.

| Variables | Policy citations |  |
| --- | --- | --- |
|  | OR (95% CI) | P value |
| **Years since publication** | **1.53(1.22-1.91)** | **<0.001** |
| JIF(2022) | 1.01 (0.99-1.03) | 0.150 |
| Number of authors | 1.01(0.98-1.03) | 0.655 |
| **Number of affiliations** | **1.14 (1.01-1.28)** | **0.029** |
| **Publication type** |  | **0.035** |
| Research article | 1(ref) |  |
| **Review** | **0.35(0.13-0.89)** | **0.029** |
| SR/MA | 2.40(0.66-8.65) | 0.180 |
| Other | 1.60(0.26-9.67) | 0.607 |
| **Paper's status** |  | **0.283** |
| Top-cited papers | 1(ref) |  |
| "Super-papers" | 1.88(0.61-5.77) | 0.270 |
| Top-AAS papers | 0.77(0.23-2.56) | 0.669 |

Abbreviations: AAS: Altmetric Attention Score; JIF: Journal Impact Factor; SR/MA: Systematic Review/Meta-Analysis; Definitions: "Super-papers" -Duplicate articles appearing in both 100 top-cited papers and 100 top-AAS papers.

#### Burden/policy papers

Among the burden/policy papers excluding duplicates (n= 159), 55 papers (34.6%) had been cited in at least one policy document. A comparison of the characteristics between the papers that were cited within policy documents versus those that were not is depicted in **Table 7**. Papers with policy citations had a significantly higher number of years since publication and a larger proportion of “super-papers”. The number of patent and Wikipedia mentions in the papers with policy citations was also significantly higher for the policy-cited papers.

**Table 7.** Burden/policy papers characteristics with and without policy citations.

| Variables | Papers without<br>policy citations<br>n= 104 | Papers with<br>policy citations<br>n=55 | P<br>Value |
| --- | --- | --- | --- |
| **Paper's and Journal's characteristics** |  |  |  |
| OA Status* | 70(67.3) | 36(65.5) | 0.860 |
| Publication Type* |  |  | 0.416 |
| Research Article | 53(51) | 32(58.2) |  |
| Review | 24(23.1) | 8(14.5) |  |
| SR/MA | 19(18.3) | 13(23.6) |  |
| Other | 8(7.7) | 2(3.6) |  |
| JIF Category in JCR* |  |  | 0.333 |
| Gastroenterology<br>and Hepatology | 80(76.9) | 45(81.8) |  |
| Nutrition and<br>Dietetics | 1(1) | 0 |  |
| Endocrinology and<br>Metabolism | 4(4.8) | 5(9.1) |  |
| Other | 18(17.3) | 5(9.1) |  |
| Q of JIF in JCR* |  |  | 0.720 |
| 1 | 77(74) | 42(76.4) |  |
| 2 | 18(17.3) | 7(12.7) |  |
| 3/4 | 9(8.7) | 6(10.9) |  |
| First author region* |  |  | 0.670 |
| America, Canada,<br>Australia, NZ | 53(51) | 29(52.7) |  |
| Europe | 24(23.1) | 15(27.3) |  |
| Asia, Middle East | 27(26) | 11(20) |  |
| Years since publication** | 4(4) | 6(4) | <0.001 |
| JIF (2022)** | 10(9.42) | 14(20.6) | 0.125 |
| No. of authors** | 7(6) | 8(8) | 0.196 |
| No. of affiliations** | 3(4) | 3(7) | 0.805 |
| Paper's status* |  |  | <0.001 |
| Top-cited papers | 38(36.5) | 21(38.2) |  |
| Top-AAS papers | 49(47.1) | 10(18.2) |  |
| "Super-papers" | 17(16.3) | 24(43.6) |  |
| **Components of the impact measures** |  |  |  |
| No. of citations** | 72(95) | 115(330) | <0.001 |
| No. of Mendeley readers** | 81.5(123) | 145(264) | <0.001 |
| AAS** | 27(44) | 45(79) | 0.122 |
| No. of Twitter(X)<br>mentions** | 26(49) | 26(82) | 0.755 |
| No. of news mentions** | 1(4) | 2(7) | 0.028 |
| No. of blog mentions** | 0(0) | 0(0) | 0.166 |
| No. of Patent mentions** | 0(0) | 0(0) | 0.001 |
| No. of Facebook<br>mentions** | 0(1) | 0(1) | 0.808 |
| No. of Wikipedia<br>mentions** | 0(0) | 0(0) | 0.010 |
| No. of YouTube mentions** | 0(0) | 0(0) | 0.899 |

\*n(%); \*\* Median (IQR); Abbreviations: AAS: Altmetric Attention Score; JCR: Journal Citation Report (Clarivate); JIF: Journal Impact Factor; No.: Number; OA: Open Access; Q: Quartile; SR/MA: Systematic Review/Meta-Analysis.

In multivariable analysis (**Table 8**) including all variables that were significantly or borderline-significantly different between cited versus uncited papers in the univariable analysis (**Table 7**), the significant predictors for being cited in policy were: number of years since publication (OR=1.33; 95% CI 1.14-1.56, P<0.001, per one-year increase), JIF (OR=1.03;95% CI 1.01-1.06 P=0.032, per one-unit increase) and being a “super-paper” versus a top-cited (OR=2.60; 95% CI 1.06-6.38, P=0.036) or top-AAS paper (OR=4.53; 95%CI 1.68-12.19, P=0.003) (calculated from the model).

**Table 8.**
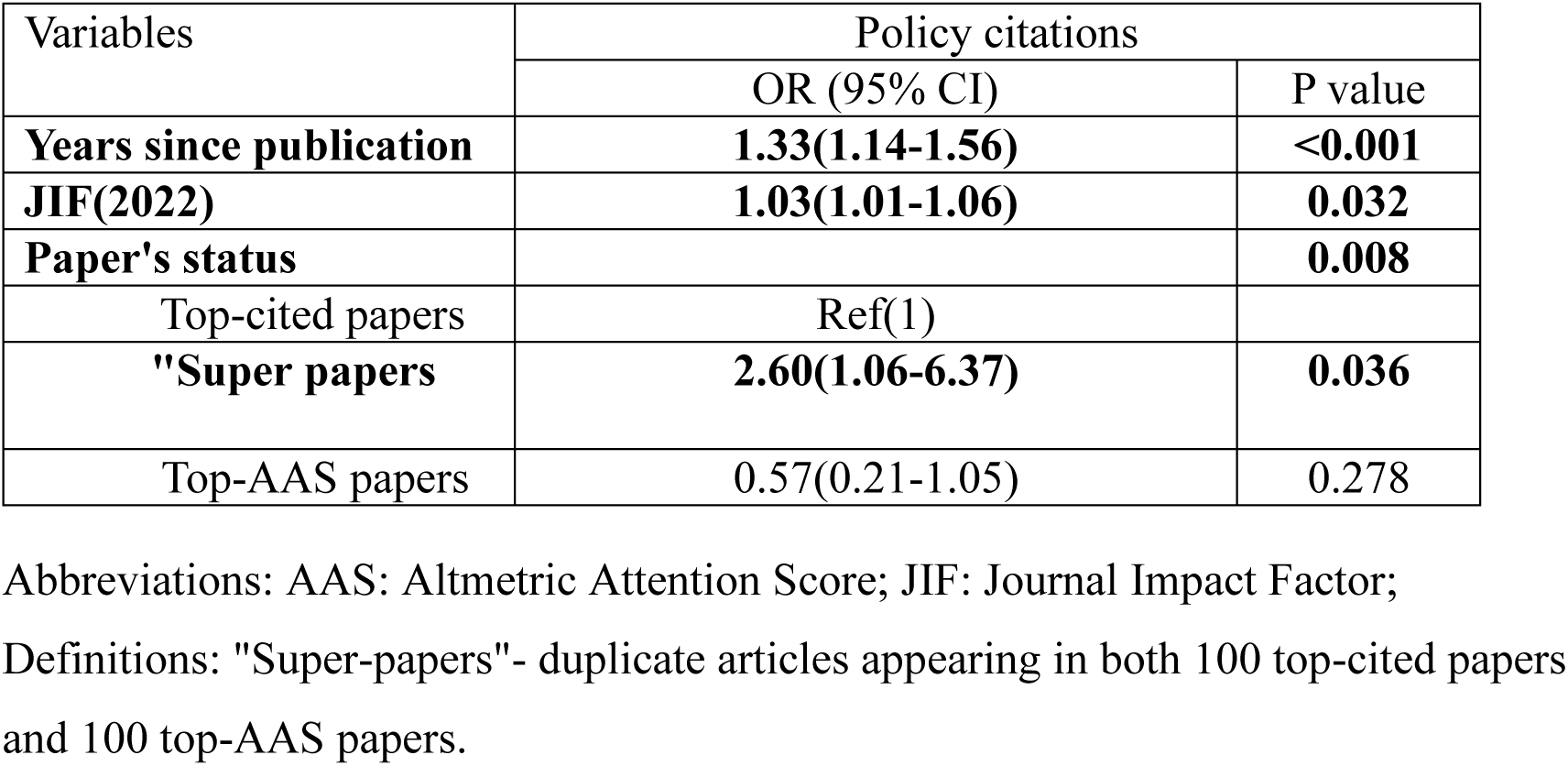
Multivariable analysis of the association between burden/policy papers characteristics and policy citations.

## Discussion

This study has explored the academic (via citation metrics) and societal impact (via AAS and policy citations metrics) of papers, and the correlation between these metrics, in two main fields of MASLD research: lifestyle and burden/policy. Over the past 20 years, papers on MASLD/MASH have grown exponentially, in line with the rising prevalence of this liver disease and emerging knowledge. However, more effort is needed to translate these publications into increased health system preparedness, policy action and public awareness (18, 19). By identifying the paper’s, journal’s, and author‘s characteristics associated with higher metrics and impact, our study may contribute to a better understanding of how to improve policy impact.

The study demonstrated a moderate positive correlation in both fields of research between citations and AAS in top-cited papers. This suggests that highly cited papers tended to also have higher AAS or that papers with higher online engagement tend to receive more citations, but the heterogeneous nature of AAS makes it hard to interpret. The moderate correlation also suggests that there are other factors affecting citation and AAS that should be explored. Several previous studies examined the correlation between citations and AAS with mixed results ranging between r of 0.18 to 0.64(20–23). The variation in methods for obtaining the research datasets, along with differences between fields of research, may account for the different results.

Furthermore, in our study, the overlap between the top-cited and top-AAS papers in each field of study (the “super-papers”) was considerably high. In contrast, for example, a study of obstetrics and gynecology papers found no overlap between the 100 top-cited and 100 top-AAS papers lists(21), however, unlike our study, the papers were not selected based on specific topics.

Interestingly, in our data, the majority of the top-AAS/cited papers in lifestyle and burden/policy fields were OA. Furthermore, the main “super-paper” predictor we found was being an open-access paper. The top-cited papers were less likely to be published in open access compared to the “super-papers” and top-AAS papers. Other studies also found that top-AAS papers had a higher proportion of open access status compared to top-cited papers (21, 24). These findings suggest that publishing open-access papers may be important for achieving greater exposure and impact beyond the academic community. However, we did not find an association between OA Status and getting policy document citations, unlike other studies (25, 26).

JIF was found as a predictor of “super-paper” status as well as of policy citations. Others have found similar results(27). It should be noted that the influence of the JIF on metrics analysis may change in the coming years due to the inclusion of all Web of Science Core Collection journals, including arts and humanities and multidisciplinary emerging sources, in the JIF ranking starting from the 2023 edition of the JCR database(28). The inclusion of new journals with different characteristics in the JIF ranking may change its impact on metrics analysis in future research.

The percentage of papers with at least one policy citation in our findings was higher (33.3% and 34.6% in the lifestyle and burden/policy fields respectively) than in previous studies. Early studies analyzing the proportion of papers with policy citations based on Altmetric data showed very low proportions of 0.3%(29) and 1.2%(30). Later studies using Overton database data showed higher proportions of 5.1-8.6%(31), 13%(27), 14%(25) and 16%(32). The higher rate of papers with policy citations in our results can be explained by the fact that we focused exclusively on the top-ranked papers in each field, whereas other studies analyzed a broader range of papers, regardless of their rank.

The strongest predictor of getting a policy citation in our study was being a “super-paper” within the burden/policy field. The lifestyle papers showed the same tendency, but it wasn’t statistically significant. A very similar work regarding the impact of physical activity research on policy also compared the 100 top-cited and 100 top-AAS papers (33). The data evaluation differed, yet the conclusion aligned in the same spirit: Influencing policy requires more than scientific publication; it demands collaborative engagement strategies with policymakers and communities.

In addition to “super-paper” status, other factors were associated with the likelihood of being cited in a policy document. The number of years since publication was significant in both fields and showed that newer papers are less likely to be cited in a policy document. Other studies found similar results(25, 27). Pinheiro et al(31) found that approximately 50% of the papers funded by certain prestigious grants got their first policy citation three years after their publication.

In the lifestyle field only, our findings showed the publication type of a research article, as opposed to a review, increases the likelihood of a paper being cited in a policy document, similar to previous studies (31). Also, the number of affiliations was associated with the likelihood of being cited in a policy document in the lifestyle field only. Other studies also found a higher likelihood of policy citations for papers with more disciplinary diversity of authors, cross-disciplinary interactions and a higher number of countries (31, 32, 34). A greater number of affiliations or countries may suggest a more interdisciplinary and global approach, which could be particularly valuable in addressing complex issues, such as those typically tackled in policy papers. In contrast, the number of authors was not related to being cited in a policy document in both fields of study. Other studies also found similar results(32) or even reduced probability of policy citations for papers with a greater number of authors(31).

“Super-papers” and the papers with policy citations compared to both top-cited and top-AAS papers or to the papers with no policy citations had more mentions on Facebook, Wikipedia and YouTube. While these findings might suggest that promoting papers in these channels can help get better paper dissemination and impact, they require further validation in future studies. Their association with “super-papers” or policy citation status could not be assessed in the multivariable analysis, as they are already factored into the AAS score. Patent mentions were also significantly higher in the “super-papers” and the papers with policy citations, which may suggest that influential papers can impact society in various ways, including innovation and industry support. Patent citations were also previously suggested as a possible metric for scientific research impact(17, 35).

The strength of our study is the meticulous methodology of data extraction from several resources. The detailed structured procedures described in this study can serve as an example of scientific impact analysis. We covered two fields of knowledge, which enabled a broader perspective on the MASLD literature and also a comparison between the fields and identifying variability in the factors predicting “super paper” status and getting a policy citation.

Another advantage of our study is using policy citation data from both AE and Overton data. More than 80% of the policy citations in our data were from the Overtone database and the duplicate proportion between AE and Overton databases was 2% only, similar to other recent studies(36, 37). These results emphasize the importance of combining data from more than one source to get more accurate analysis. Future studies may use also another new database of citations of papers in international clinical practice guidelines, named Clinical Impact (38).

There are several limitations to consider in our study and future perspectives. The study was field-specific, reducing the generalizability of the findings to other fields. Regarding the author’s region, we analyzed the data based on the region of the first author. However, analyzing the region of the last author might yield different results, as the last author is often the senior researcher, who typically has greater influence on the research topics and budget allocation. The data on citations and altmetrics are dispersed across several databases, making it more challenging to collect, and increasing the potential for errors. Additionally, Altmetric Explorer, the major database for comprehensive altmetrics research, is not designed for complex searches. The only way available at this point to build a complex search in AE is via a PubMed query, which is limited to 10,000 results, and not useful for research in fields other than medicine. These search limitations may result in missing relevant data for research.

Future research may establish a consensus on the definition of policy documents and conduct a stratified analysis of the data based on different types of policy documents, which may yield varying results. Other directions for future research include exploring paper citations in patents and Wikipedia as a metric of research impact in separation from the heterogenous AAS and investigating the effect of different types of open access papers—such as gold, green, and bronze—on paper’s impact. Moreover, conducting similar studies in additional fields within gastroenterology and hepatology, in other fields in medicine and beyond would help determine whether our findings are consistent or if variations exist across different disciplines.

In conclusion, our study found that papers with high metrics (“super-papers”), together with additional factors such as JIF, years since publication and number of affiliations, make more impact as measured by citations in policy documents. This information can help researchers plan their paper’s publishing and dissemination processes and consider new dissemination mediums to contribute to MASLD/MASH policy-making.

## Data Availability

All data produced are available online at https://zenodo.org/records/16315405

## List of abbreviations

AAS: Altmetric Attention Score
AE: Altmetric Explorer
AI: artificial intelligence
CI: confidence interval
CPG: clinical practice guideline
DOI: digital object identifier
IQR: interquartile range
JCR: Journal Citation Report
JIF: Journal Impact Factor
MASH: metabolic dysfunction-associated steatohepatitis
MASLD: metabolic dysfunction-associated steatotic liver disease
NAFLD: non-alcoholic fatty liver disease
NASH: non-alcoholic steatohepatitis
OA: open access
OR: odds ratio
PMC: PubMed Central
PRISMA: Preferred Reporting Items for Systematic reviews and Meta-Analyses
PP: proceeding paper
Q: quartile
r_s_: Spearman’s correlation coefficient
SD: standard deviation
SR/MA: systematic review/meta-analysis
TLVMC: Tel-Aviv Medical Center
WoS: Web of Science

## Acknowledgments

The authors wish to thank Altmetric and Overton for providing this study’s data free of charge for research purposes.

JVL acknowledges support to ISGlobal from grant CEX2023-0001290-S, funded by MCIN/AEI/ 10.13039/501100011033, and support from the Generalitat de Catalunya, through

## Disclosure of Ethical Statements

Approval of the research protocol: N/A

Informed Consent: N/A

Registry and the Registration No. of the study/trial: N/A

Animal Studies: N/A

Research involving recombinant DNA: N/A

## Data Availability Statement

The data associated with this article is available in Zenodo at https://zenodo.org/records/16315405

## Appendices

## Appendix I. Study Variables

The variables in the study are divided into paper’s level variables, author’s level variables and journal’s level variables.

### Paper’s level variables

#### Number of Citations

the number of citations for each paper was extracted from WOS platform all-databases data (Clarivate). This is a continuous variable, serving as a dependent variable in relation to the top-cited papers and as an independent variable in relation to the top-AAS papers.

#### Altmetric Attention Score (AAS)

AAS is an automatically calculated weighted count of all the attention a research output has received online, in sources tracked by Altmetric company (https://help.altmetric.com/support/solutions/articles/6000233311-how-is-the-altmetric-attention-score-calculated-). The AAS data was extracted from the Altmetric Explorer (AE) database. This data represents a continuous variable and serves as a dependent variable when analyzing the top-AAS papers, while it acts as an independent variable when examining the top-cited papers.

#### Publication Type

A nominal independent variable indicating the kind of paper published. Publication types included: research articles (Article), narrative reviews (Review), systematic review/meta-analysis(SR/MA), other types of publications (grouped as OTHER): proceeding paper (PP), comment, editorial, clinical practice guidelines, consensus papers.

#### Open Access Status (OA Status)

A dichotomous independent variable indicating whether the paper is freely accessible or requires a subscription/payment for access.

#### Years Since Publication

A continuous potential confounder variable representing the number of years since the paper was first published (as indicated in the WOS platform or AE database) up to 2024.

#### Number of Policy Citations

An independent continuous variable representing the total number of unique policy documents in which the paper is mentioned, as determined by data from the Altmetric Explorer and Overton databases.

#### Number of Patent Mentions

A continuous independent variable representing the number of patents mentions of the paper as listed in Altmetric Explorer database.

#### Number of Twitter (x) Mentions

A continuous independent variable representing the number of Twitter(x) mentions of the paper as listed in Altmetric Explorer database. The total number of posts was retrieved, without filtering posts from the same user account.

#### Number of Mendeley Readers

A continuous independent variable representing the number of Mendeley Readers of the paper as listed in Altmetric Explorer database.

#### Number of News Mentions

A continuous independent variable representing the number of news mentions of the paper as listed in Altmetric Explorer database. The total number of posts was retrieved, without filtering posts from the same user account.

#### Number of Blog Mentions

A continuous independent variable representing the number of blog mentions of the paper as listed in Altmetric Explorer database. The total number of posts was retrieved, without filtering posts from the same user account.

#### Number of Facebook Mentions

A continuous independent variable representing the number of Facebook mentions of the paper as listed in Altmetric Explorer database. The total number of posts was retrieved, without filtering posts from the same user account.

#### Number of Wikipedia Mentions

A continuous independent variable representing the number of Wikipedia mentions of the paper as listed in Altmetric Explorer database. The total number of posts was retrieved, without filtering posts from the same user account.

#### Number of YouTube Mentions

A continuous independent variable representing the number of YouTube mentions of the paper as listed in Altmetric Explorer database. The total number of posts was retrieved, without filtering posts from the same user account.

#### Paper’s status variable

Independent nominal variable grouping the papers in each field based on their classification: those falling into the top-AAS papers only category, those in the to-cited papers only category, and those belonging to both categories, referred to as duplicates or “super-papers”.

### Author’s level variables

#### Number of Authors

A continuous independent variable representing the total count of individuals who contributed to the writing of the paper, as listed in the published paper in the Web of Science (WOS) platform.

#### Number of Affiliations

A continuous independent variable representing the number of distinct institutional affiliations listed for the paper’s authors in WOS, with different departments or faculties within the same institution counted as one.

#### First Author Region

A nominal independent variable representing the region listed for the first author in the published paper in WOS platform. The regions were grouped as follows: (1) North & South America, Canada, Australia& NZ (2)Europe, (3)Asia & Middle East.

### Journal’s level variables

#### Journal Impact Factor (JIF)

A continuous independent variable representing the JIF score of the journal the paper was published in from JCR database (Clarivate) 2022 edition.

#### Journal Category in JCR

A nominal independent variable indicating the category of the journal in which the paper was published as listed in the 2022 edition of the Journal Citation Report (JCR) database (Clarivate). If the journal was listed in more than one category, the category with the higher quartile (Q) ranking was chosen. If the quartiles ranking was identical for 2 or more categories, the category most closely related to the paper’s subject was selected. The categories are as follows: (1)Gastroenterology & Hepatology, (2)Nutrition & Dietetics, (3)Endocrinology & Metabolism, (4)Other. The “Other” grouping include the following categories: Pediatrics, Sport Sciences, Medicine General & Internal, Medicine Research & Experimental, Multidisciplinary Sciences, Immunology, Biochemistry & Molecular Biology, Toxicology, Physiology, Transplantation, Health Policy & Services, Food Science & Technology, Gerontology, Pharmacology & Pharmacy, Cardiac & Cardiovascular Systems, Surgery.

#### Quartile(Q) of JIF in JCR category

An ordinal independent variable representing the Quartile (Q) of the JIF in the journal’s category as listed in JCR database (Clarivate) 2022 edition. The quartiles were grouped into 3 categories: the original quartiles 1 and 2 as separate categories, and quartiles 3 and 4 combined into one category (3/4).

## Appendix II. Comparative analysis of the coverage of policy-related data in the Overton and Altmetric Explorer databases

| Papers group | Total policy citations | No. of AE policy citations (% of total) | No. of Overton policy citations (% of total) | No. of policy citations duplicates (% of total) | Total policy citations (no duplicates) |
| --- | --- | --- | --- | --- | --- |
| Burden/policy papers (no duplicates)<br>n=159 | 113 | 22(19.4%) | 91(80.5) | 2 (2%) | 111 |
| Lifestyle papers (no duplicates)<br>n=174 | 153 | 19(12.4%) | 134(87.5) | 3 (2%) | 150 |

## Appendix III. Artificial Intelligence tools use

### Affiliation duplicate master tools and prompt

Affiliations Duplicate Master: During the data extraction for each paper the number of affiliations of the authors was recorded from WOS platform. We considered different departments in the same institution as one affiliation. When a paper had many authors, or the authors had many different affiliations, an AI tool was used to help get accurate results.

The tools used for finding affiliations duplicates were Perplexity AI and Cloud 3 (free versions). The prompt used: “I will give you a list of authors of an academic article with different affiliations. Please make a table with a column counting how many authors are from the same affiliation. Group the authors from the same institution regardless of their department. In the end sum the total number of different affiliations. Do not search for information about these institutions.” Each result generated by the tools was cross-verified against the data.

### Julius AI prompt and code for de-duplication

De-duplication of papers in each field of study was done with the help of the tool Julius AI paid version. The de-duplication of papers for each field of study was done according to the DOI number of the papers. It was done on 25.5.24 using the tool Julius.ai available on https://julius.ai/. The process was repeated 3 times for each field, and then double-checked for each duplicate DOI.

#### The prompts used

1. According to the DOI, are there any duplicates across the 2 lifestyle files? create a table with the results.
2. Repeat with the 2 burden files.

#### Python code generated for lifestyle papers

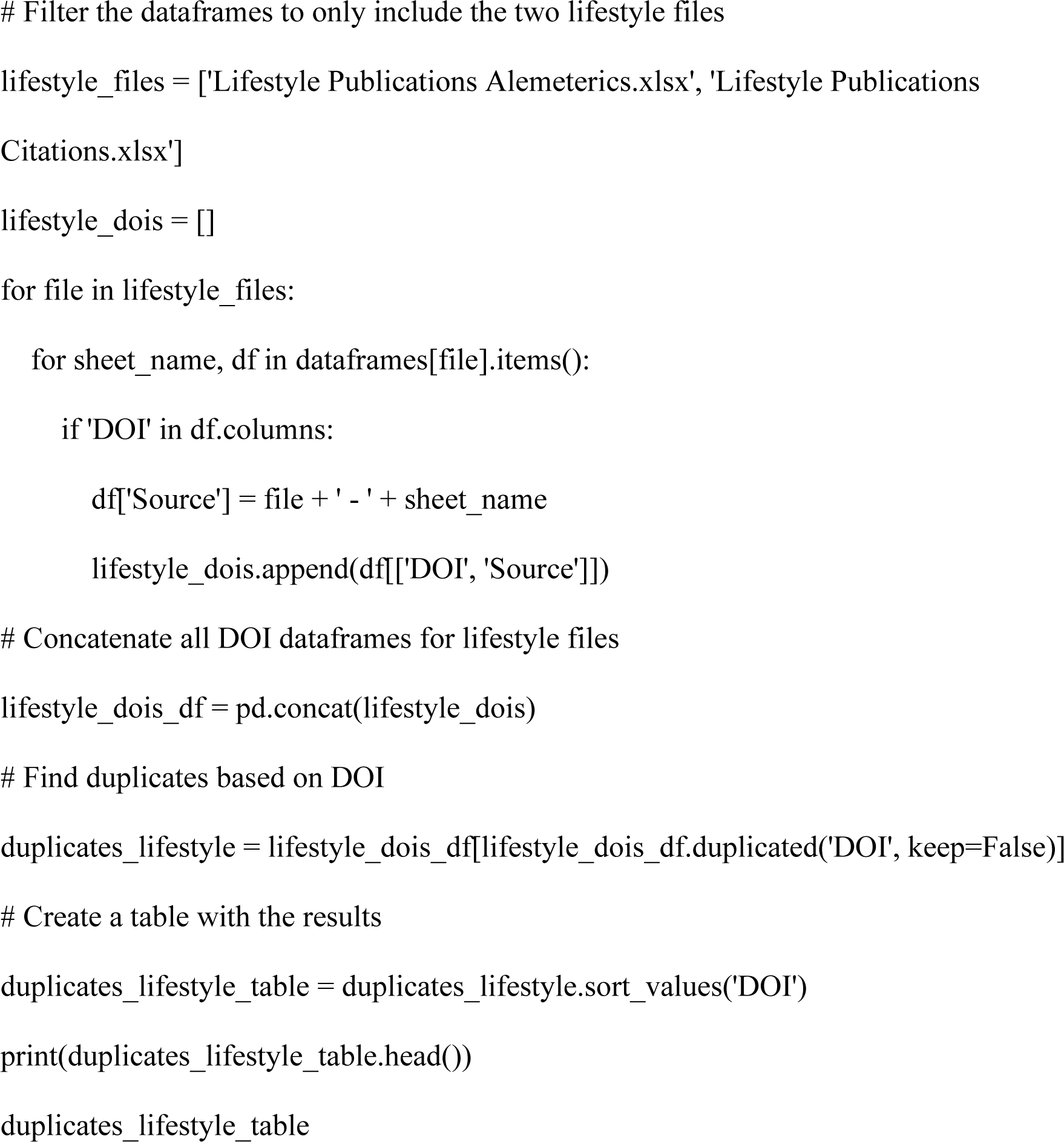

#### Python code generated for burden/policy papers

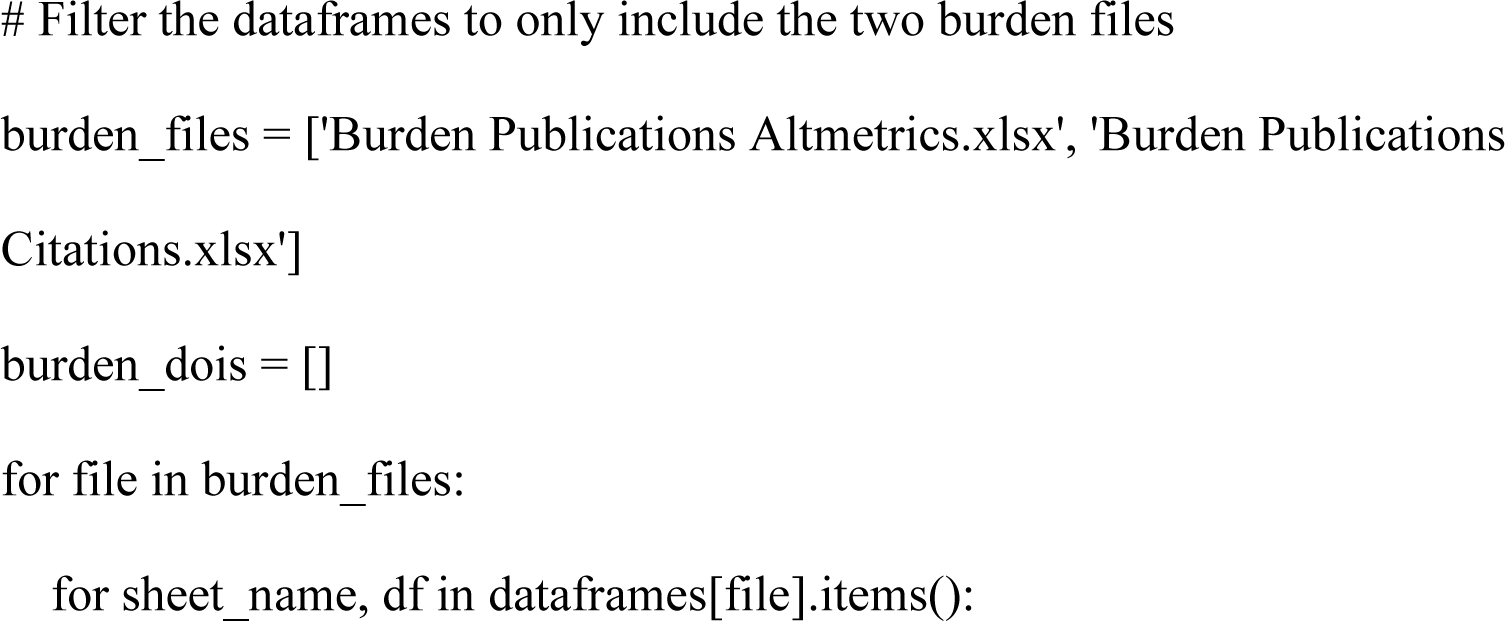
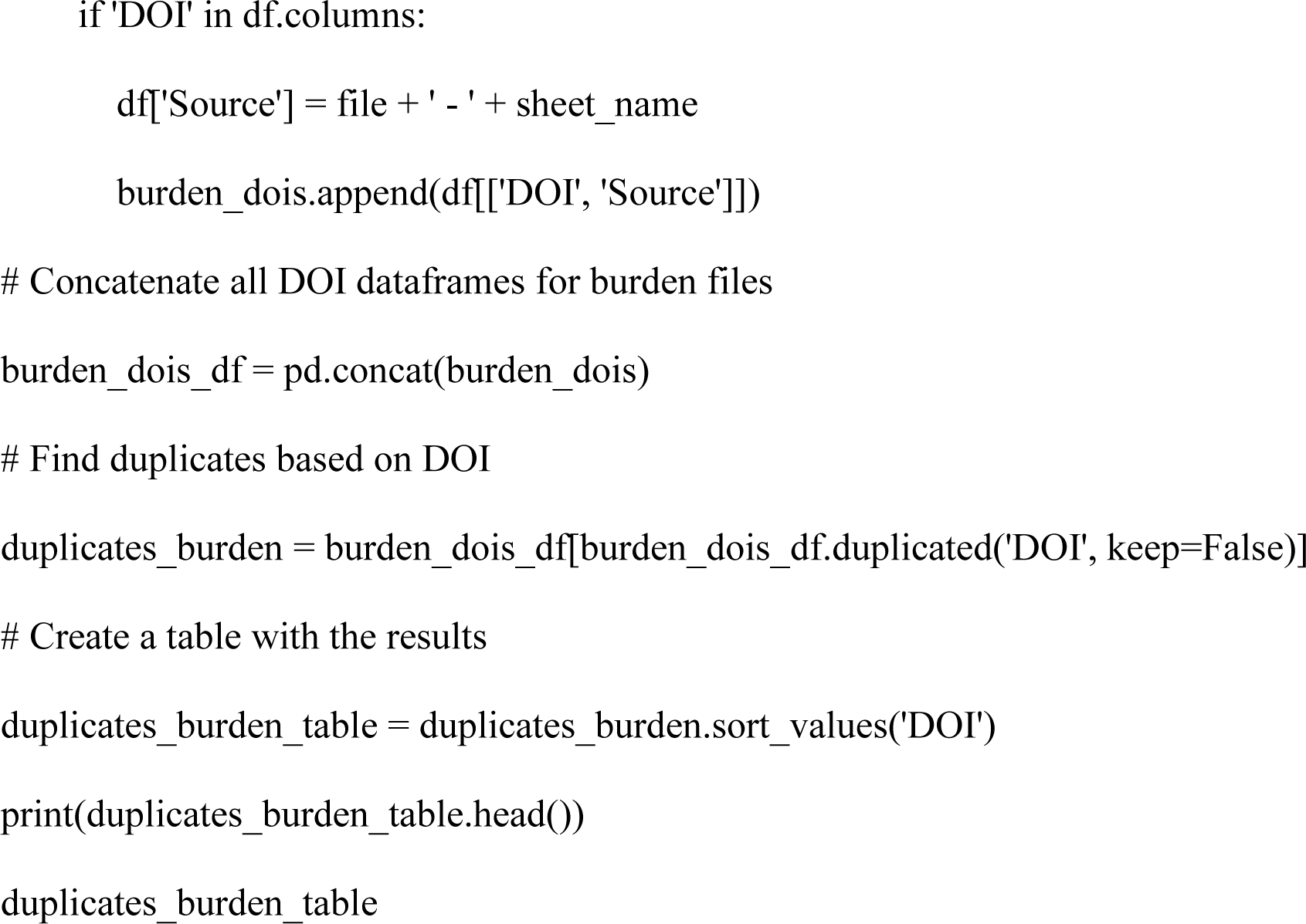

### Paraphrasing authors own ideas

Paraphrasing authors own ideas for clearer and academic suitable style ChatGPT4o free version was used.

