## Supplemental online material for "“Super-Papers” Make More Impact: A Bibliometric and Altmetric Databases Analysis of MASLD Publications"

### Supplemental online materials

#### Supplement 1. Web of Science search strategy for lifestyle papers

Day of search: 15.01.2024

**Databases searched in WOS platform**: WOS: 1965 to 2024, KJD: 1980 to 2024, MEDLINE: 1950 to 2024, PPRN: 1991 to 2024, PQDT: 1637 to 2024, SCIELO: 2002 to 2024, ZOOREC: 1995 to 2024.

| # | Search Query | Results |
|---|---|---|
| 1 | TI=('metabolic liver disease' OR 'metabolic associated liver disease ' OR 'Metabolic dysfunction-associated fatty liver disease' OR mafld) OR AB=('metabolic liver disease' OR 'metabolic associated liver disease ' OR 'Metabolic dysfunction-associated fatty liver disease' OR mafld) | 35357 |
| 2 | TI=('nonalcoholic steatotic hepatitis' OR 'nonalcoholic steatosis hepatitis') OR AB=('nonalcoholic steatotic hepatitis' OR 'nonalcoholic steatosis hepatitis') | 585 |
| 3 | TI=('nonalcohol steato-hepatitis' OR 'nonalcohol steatohepatitis') OR AB=('nonalcohol steato-hepatitis' OR 'nonalcohol steatohepatitis') | 17 |
| 4 | TI=('non alcoholic steato-hepatitis' OR 'non-alcoholic steatohepatitis' OR 'non-alcoholic steatosis hepatitis' OR 'non-alcoholic steatotic hepatitis') | 4269 |
| 5 | TI=('non alcohol steato-hepatitis' OR 'non alcohol steatohepatitis') | 33 |
| 6 | TI=(NASH OR MASH OR 'metabolic dysfunction associated steatohepatitis') OR AB=( MASH OR 'metabolic dysfunction associated steatohepatitis') | 22435 |
| 7 | TI=('nonalcoholic hepatosteatosis' OR 'nonalcoholic liver steatosis') OR AB=('nonalcoholic hepatosteatosis' OR 'nonalcoholic liver steatosis') | 7306 |
| 8 | TI=('non alcoholic steatotic hepatopathy ' OR 'non-alcoholic FLD' OR 'non-alcoholic hepatic steatosis' OR 'nonalcoholic FLD' OR 'nonalcoholic hepatic steatosis') OR AB=('non alcoholic steatotic hepatopathy ' OR 'non-alcoholic FLD' OR 'non-alcoholic | 9547 |

| | | |
|---|---|---|
| | hepatic steatosis' OR 'nonalcoholic FLD' OR 'nonalcoholic hepatic steatosis') | |
| 9 | TI=('non alcoholic hepato steatosis' OR 'non alcoholic hepatosteatosis' OR 'non alcoholic liver steatosis') OR AB=('non alcoholic hepato steatosis' OR 'non alcoholic hepatosteatosis' OR 'non alcoholic liver steatosis') | 7355 |
| 10 | TI=( 'metabolic dysfunction-associated steatotic liver disease' OR MASLD) OR AB=( 'metabolic dysfunction-associated steatotic liver disease' OR MASLD) | 341 |
| 11 | TI=(nafld OR 'nonalcoholic fatty liver disease' OR 'non alcoholic fatty liver disease' ) OR AB=(nafld OR 'nonalcoholic fatty liver disease' OR 'non alcoholic fatty liver disease' ) | 47654 |
| 12 | TI=('fatty liver') OR AB=('fatty liver') | 108056 |
| 13 | #1 OR #2 OR #3 OR #4 OR #5 OR #6 OR #7 OR #8 OR #9 OR #10 OR #11 OR #12 | 152601 |
| 14 | TI=(weight OR body-mass ) | 384520 |
| 15 | TI=(nutr* OR diet*) | 776512 |
| 16 | TI=(fat OR food* OR calori* OR sugar* OR fructose OR fatty-acid* OR dietary-fiber* OR corn-syrup) | 877523 |
| 17 | TI=('Healthy Eating Index' OR 'advanced glycation' OR ' advanced glycosylation' OR Maillard) | 12205 |
| 18 | TI=(Life-Style OR lifestyle OR exercis* OR 'physical activity' OR Sport* OR training OR Behavior* OR 'Risk Reduction') | 1967846 |
| 19 | TI=(meat* OR poultry OR fish OR nut* OR grain* OR legume* OR vegetable*OR fruit* OR dairy OR butter OR yogurt OR oil* OR cholesterol) | 1389442 |
| 20 | TI=(salt OR sodium OR glucose OR sucrose OR Isoglucose OR Maize-Syrup OR beverage* OR soda*OR drink*OR juice*) | 710178 |
| 21 | TI=(liquor* OR coffee OR tea* OR eat*) | 833510 |
| 22 | TI=(treatment* OR management OR therap*) | 4661664 |
| 23 | #14 OR #15 OR #16 OR #17 OR #18 OR #19 OR #20 OR #21 OR #22 | 10570189 |

| | | |
|---|---|---|
| 24 | #13 AND #23 | 50406 |
| 25 | #13 AND #23 and ProQuest™ Dissertations & Theses Citation Index or Zoological Record or Preprint Citation Index (Exclude – Database) | 47746 |
| 26 | #13 AND #23 and ProQuest™ Dissertations & Theses Citation Index or Zoological Record or Preprint Citation Index (Exclude – Database) and 2024 or 2023 or 2022 or 2021 or 2020 or 2014 or 2015 or 2016 or 2017 or 2018 or 2019 (Publication Years) | 23960 |

**Supplement 2. Altmetric search strategy for lifestyle papers**

Day of search: 15.01.2024

**Search strategy:**

(("treatment*"[Title] OR "management"[Title] OR "therap*"[Title] OR "coffee"[Title] OR "tea"[Title] OR "eat"[Title] OR "salt"[Title] OR "sodium"[Title] OR "glucose"[Title] OR "sucrose"[Title] OR "Isoglucose"[Title] OR "Maize-Syrup"[Title] OR "beverage*"[Title] OR "soda*"[All Fields] OR "drink*"[All Fields] OR "juice*"[Title] OR "meat*"[Title] OR "poultry"[Title] OR "fish"[Title] OR "nut"[Title] OR "nuts"[Title] OR "grain*"[Title] OR "legume*"[Title] OR "vegetable*"[Title] OR "fruit*"[Title] OR "dairy"[Title] OR "butter"[Title] OR "yogurt"[Title] OR "oil"[Title] OR "oils"[Title] OR "cholesterol"[Title] OR "Life-Style"[Title] OR "lifestyle"[Title] OR "exercis*"[Title] OR "training"[Title] OR "physical activity"[Title] OR "sport*"[Title] OR "behavior*"[Title] OR "risk reduction"[Title] OR "healthy eating index"[Title] OR "advanced glycation"[Title] OR "advanced glycosylation"[Title] OR "Maillard"[Title] OR "fat"[Title] OR "food*"[Title] OR "calori*"[Title] OR "sugar*"[Title] OR "fructose"[Title] OR "fatty acid*"[Title] OR "dietary fiber*"[Title] OR "corn-syrup"[Title] OR "nutr*"[Title] OR "diet*"[Title] OR "weight"[Title] OR "body-mass"[Title]) AND ("metabolic liver disease"[tiab] OR "metabolic associated liver disease"[ tiab] OR "nonalcoholic steato-hepatitis"[ tiab] OR "nonalcoholic steatohepatitis"[ tiab] OR "non-alcoholic steato-hepatitis"[ tiab] OR "non-alcoholic steatohepatitis"[ tiab] OR "non alcohol steatohepatitis"[ tiab] OR "NASH"[Title] OR "nonalcoholic hepatosteatosis"[tiab] OR "nonalcoholic liver steatosis"[tiab] OR "non-alcoholic hepatic steatosis"[ tiab] OR "non-alcoholic hepatic steatosis"[ tiab] OR "non alcoholic hepatosteatosis"[tiab] OR "non alcoholic liver steatosis"[tiab] OR "mafld"[ tiab] OR "Metabolic dysfunction-associated fatty liver disease"[ tiab] OR "nafld"[ tiab] OR "fatty-liver"[ tiab] OR "non alcoholic fatty liver disease"[ tiab] OR "nonalcoholic fatty liver disease"[tiab] OR "metabolic dysfunction-associated steatotic liver disease"[ tiab] OR "MASLD"[ tiab] OR MASH [tiab] OR "metabolic dysfunction associated steatohepatitis"[tiab])) AND (2014/1/1:2030/1/1[pdat])

**Supplement 3. Web of Science search strategy for burden/policy papers**

Date of search: 07.02.2024

**Databases searched in WOS platform**: WOS: 1965 to 2024, KJD: 1980 to 2024, MEDLINE: 1950 to 2024, PPRN: 1991 to 2024, PQDT: 1637 to 2024, SCIELO: 2002 to 2024, ZOOREC: 1995 to 2024.

| # | Search Query | Results |
|---|---|---|
| 1 | TI=(model*) | 3552095 |
| 2 | TI=(capital OR expenditure* OR access* OR resource* OR service*) | 1333429 |
| 3 | TI=('Community Health' OR 'international health' OR 'national health' OR 'World Health' OR 'Worldwide Health' OR 'World wide Health' OR 'Care Map*') | 137609 |
| 4 | TI=(leader* OR Influential* OR Theor* OR 'health system*') | 1408100 |
| 5 | TI=(plans OR planning OR 'public health' OR epidem* OR 'health service*' OR 'social determinants of health' OR Awareness ) | 976660 |
| 6 | TI=(goal* OR Sustainab* OR burden* OR polic* OR 'health care' OR healthcare OR cost* OR economic*) | 2135723 |
| 7 | TI=(prevalence OR incidence) | 465659 |
| 8 | #1 OR #2 OR #3 OR #4 OR #5 OR #6 OR #7 | 9305512 |
| 9 | TI=('metabolic liver disease' OR 'metabolic associated liver disease ' OR 'Metabolic dysfunction-associated fatty liver disease' OR mafld) OR AB=('metabolic liver disease' OR 'metabolic associated liver disease ' OR 'Metabolic dysfunction-associated fatty liver disease' OR mafld) | 38229 |
| 10 | TI=('nonalcoholic steatotic hepatitis' OR 'nonalcoholic steatosis hepatitis') OR AB=('nonalcoholic steatotic hepatitis' OR 'nonalcoholic steatosis hepatitis') | 604 |
| 11 | TI=('nonalcohol steato-hepatitis' OR 'nonalcohol steatohepatitis') OR AB=('nonalcohol steato-hepatitis' OR 'nonalcohol steatohepatitis') | 17 |
| 12 | TI=('non alcoholic steato-hepatitis' OR 'non-alcoholic steatohepatitis' OR 'non-alcoholic steatosis hepatitis' OR 'non-alcoholic steatotic hepatitis') | 4557 |
| 13 | TI=('non alcohol steato-hepatitis' OR 'non alcohol steatohepatitis') | 32 |

| | | |
|---|---|---|
| 14 | TI=(NASH OR MASH OR 'metabolic dysfunction associated steatohepatitis') OR AB=( MASH OR 'metabolic dysfunction associated steatohepatitis') | 23307 |
| 15 | TI=('nonalcoholic hepatosteatosis' OR 'nonalcoholic liver steatosis') OR AB=('nonalcoholic hepatosteatosis' OR 'nonalcoholic liver steatosis') | 7588 |
| 16 | TI=('non alcoholic steatotic hepatopathy ' OR 'non-alcoholic FLD' OR 'non-alcoholic hepatic steatosis' OR 'nonalcoholic FLD' OR 'nonalcoholic hepatic steatosis') OR AB=('non alcoholic steatotic hepatopathy ' OR 'non-alcoholic FLD' OR 'non-alcoholic hepatic steatosis' OR 'nonalcoholic FLD' OR 'nonalcoholic hepatic steatosis') | 10034 |
| 17 | TI=('non alcoholic hepato steatosis' OR 'non alcoholic hepatosteatosis' OR 'non alcoholic liver steatosis') OR AB=('non alcoholic hepato steatosis' OR 'non alcoholic hepatosteatosis' OR 'non alcoholic liver steatosis') | 7730 |
| 18 | TI=( 'metabolic dysfunction-associated steatotic liver disease' OR MASLD) OR AB=( 'metabolic dysfunction-associated steatotic liver disease' OR MASLD) | 437 |
| 19 | TI=(nafld OR 'nonalcoholic fatty liver disease' OR 'non alcoholic fatty liver disease' ) OR AB=(nafld OR 'nonalcoholic fatty liver disease' OR 'non alcoholic fatty liver disease' ) | 50501 |
| 20 | TI=('fatty liver') OR AB=('fatty liver') | 112882 |
| 21 | #9 OR #10 OR #11 OR #12 OR #13 OR #14 OR #15 OR #16 OR #17 OR #18 OR #19 OR #20 | 159976 |
| 22 | #8 AND #21 | 11484 |

**Supplement 4. Altmetric search strategy for burden/policy papers**

Date of search: 07.2.24

**Search strategy:**

(("model*"[Title] OR "capital"[Title] OR "expenditure*"[Title] OR "access*"[Title] OR "resource*"[Title] OR "service*"[Title] OR "community health"[Title] OR "international health"[Title] OR "national health"[Title] OR "world health"[Title] OR "worldwide health"[Title] OR "world wide health"[Title] OR "care map*"[Title] OR "leader*"[Title] OR "influential*"[Title] OR "theor*"[Title] OR "health system*"[Title] OR "plan*"[Title] OR "public health"[Title] OR "epidem*"[Title] OR "health service*"[Title] OR "social determinants of health"[Title] OR "goal*"[Title] OR "sustainab*"[Title] OR "burden*"[Title] OR "polic*"[Title] OR "health care"[Title] OR "healthcare"[Title] OR "cost*"[Title] OR "economic*"[Title] OR "prevalence"[Title] OR "incidence"[Title] OR awareness[Title]) AND ("metabolic liver disease"[tiab] OR "metabolic associated liver disease"[ tiab] OR "nonalcoholic steato-hepatitis"[ tiab] OR "nonalcoholic steatohepatitis"[ tiab] OR "non-alcoholic steato-hepatitis"[ tiab] OR "non-alcoholic steatohepatitis"[ tiab] OR "non alcohol steatohepatitis"[ tiab] OR "NASH"[Title] OR "nonalcoholic hepatosteatosis"[tiab] OR "nonalcoholic liver steatosis"[tiab] OR "non-alcoholic hepatic steatosis"[ tiab] OR "non-alcoholic hepatic steatosis"[ tiab] OR "non alcoholic hepatosteatosis"[tiab] OR "non alcoholic liver steatosis"[tiab] OR "mafld"[ tiab] OR "Metabolic dysfunction-associated fatty liver disease"[ tiab] OR "nafld"[ tiab] OR "fatty-liver"[ tiab] OR "non alcoholic fatty liver disease"[ tiab] OR "nonalcoholic fatty liver disease"[tiab] OR "metabolic dysfunction-associated steatotic liver disease"[ tiab] OR "MASLD"[ tiab] OR MASH [tiab] OR "metabolic dysfunction associated steatohepatitis"[tiab])) AND (2014/1/1:2030/1/1[pdat])

**Supplement 5. Full list of 100 top-cited lifestyle papers**

The full list of 100 top-cited lifestyle papers is available at https://zenodo.org/records/16315405

**Supplement 6. Full list of 100 top-AAS lifestyle papers**

The full list of 100 top-AAS lifestyle papers is available at https://zenodo.org/records/16315405

**Supplement 7. Full list of 100 top-cited burden/policy papers**

The full list of 100 top-cited burden/policy papers is available at https://zenodo.org/records/16315405

**Supplement 8. Full list of 100 top-AAS burden/policy papers**

The full list of 100 top-AAS burden/policy papers is available at https://zenodo.org/records/16315405
